# Identification of biomedical entities from multiple repositories using a specialized metadata schema and search-augmented large language models

**DOI:** 10.1101/2025.10.21.25338460

**Authors:** Klaus Kaier, Felix Engel, Gita Benadi, Claudia Giuliani, Manuel Watter, Aref Kalantari, Karin Schuller, Claus-Werner Franzke, Markus Sperandio, Harald Binder

## Abstract

**Objective:** Many biomedical articles reference multiple datasets across different public repositories, complicating accurate metadata capture and downstream re‐use. Building on our prior grounded large language model (LLM) workflows for biomedical entity annotation, we extend the approach to identify and annotate all datasets referenced by a paper, even when distributed across repositories, by combining a specialized metadata schema with a three‐step, search‐augmented prompting strategy.

**Results:** In the Transregional Collaborative Research Center PILOT (TRR 359 “Perinatal Development of Immune Cell Topology”), Gene Expression Omnibus (GEO) releases are common alongside additional repository deposits. The applied approach reliably detected datasets referenced in articles and produced schema‐compliant annotations using information available on the repository landing pages. After validation through structured face-to-face interviews with the article’s senior author, Gemini 2.5 Pro achieved higher precision (97.1%) than GPT‐4.1 (81.9%, p<0.001) and Claude Sonnet 4 (88.6%, p<0.001). Limiting the annotation to the information available in the repositories achieved higher precision than adding information from the article (91.9 % vs. 88.3% across all LLMs, p=0.004). These results indicate that simple repository‐grounded extraction enables high quality, multi‐dataset metadata annotation which has the potential to minimize the time and effort required for manual metadata annotation.

## Introduction

Long-term research centers, such as Germany’s Collaborative Research Centers (CRCs), depend on structured data sharing and interoperable metadata to support FAIR-compliant reuse [1, 2]. In biomedicine, tagging entities like organisms, cell lines, or genes enhances data discoverability. Since manual annotation is slow and labor-intensive, automated methods such as named entity recognition are used (e.g., classifying “mouse” as an “organism”) [3]. The number and type of entities to extract depend on context and downstream analysis, varying with experimental focus (e.g., cell lines matter in in-vitro assays but not in population studies). Thus, entity selection must adapt to the task. For the CRC PILOT, which we consider as an exemplary setting, we developed a metadata schema with domain experts [4], previously used for manual annotation [5].

Many research articles now require to publish underlying datasets and more than one dataset are often distributed across multiple repositories (e.g., primary and secondary accessions, derived resources), which hinders comprehensive indexing, discovery, and integration. Prior to this work, we developed grounded large language model (LLM) workflows that combine a domain metadata schema with tool-use/validation to improve biomedical entity identification from research articles [6, 7].

Here, we address the specific challenge of automatically annotating multiple datasets per article. This fragmentation mirrors challenges seen in multi-institutional projects, where centralized, interoperable metadata platforms are proposed to unify discovery-level fields while linking discipline-specific standards across repositories [8]. We introduce a three-step prompting approach that first enumerates dataset deposits per article, then anchors annotations to repository landing pages, and finally (optionally) enriches fields using the article when repository metadata are incomplete. The method is tailored to the PILOT context, where GEO submissions are frequent but additional deposits into other archives also occur. To assess the validity of this approach, a supervised double-checking process was conducted using face-to-face interviews.

## Methods

### Prior workflow and setting

Our earlier studies evaluated grounded LLMs for biomedical entity identification using a schema-aware, multi-step generation and validation design, demonstrating high precision after expert verification [6, 7]. The present study builds on that foundation but shifts the primary evidence source from articles to repository landing pages to enhance faithfulness and reduce hallucinations, adding logic to support multiple datasets per manuscript.

### Metadata schema

We use a preliminary version of the PILOT metadata schema to standardize entities spanning, for example, *organism, timeline, cell line, tissue source, interventions, sample preparation/processing*, and *readout*, among others (controlled lists maintained with the project). The schema includes biomedical categories (e.g., *readout* and *sample processing*) and is designed for manual annotation by the responsible scientist [5]. The schema has a hierarchical structure with specific levels of data items triggering additional, more refined data items. For example, selecting ‘blood’ with the data item ‘tissue source’ would require an additional entry for data item ‘blood’ which offers a selection of blood components. As with this example, the names of data levels and data items often coincide.

### Three‐step, search‐augmented prompting

We operationalize multi‐dataset identification and annotation via a three‐step prompting approach:

1. Identify datasets in public repositories. From the manuscript and its references, enumerate all dataset deposits and capture stable URLs/identifiers (e.g., GEO, SRA, ArrayExpress, PRIDE).
2. Extract biomedical entities from each repository page. Visit each dataset’s landing page and populate schema fields exclusively from the repository record (primary source of truth).
3. Enrich from the manuscript. When repository fields are incomplete, selectively consult the article to fill missing entries.

See Supplemental Figure 1 for details on the prompting used in the three‐step approach. For each manuscript, this three -step approach was repeated for three LLMs: GPT-4.1 (2025-04-14), Gemini 2.5 Pro (2025-06-17) and Claude Sonnet 4 (2025-05-22). All models were addressed via API calls using OpenRouter. To encourage diverse and exploratory outputs from the language models, we used a temperature of 1, top-p of 1, and top-k of 0 for each LLM. The search-augmentation was implemented using the OpenRouter system for web search via API. Each model received identical step prompts, schema definitions, and retrieval instructions. Finally, the results of step 2 and step 3 were evaluated separately:

Step 2: Repository‐only content: biomedical entities were identified from the by repository pages only.

Step 3: Repository + article text: biomedical entities were identified from the by repository pages and additionally enriched using article information.

### Evaluation

Primary endpoint was precision of schema-field annotations at the dataset level. Annotation precision was validated using face-to-face interviews between a research data management expert and the article’s senior author. Scientists reviewed each annotation against their datasets and, if needed, checked methods or results sections. Suspected inaccuracies were checked against the article’s methods or results, but annotations could also be rejected based on obvious errors or doubt without comprehensive cross-checking to maintain efficiency. To estimate overall LLM accuracy, a meta-analysis of single proportions was conducted [9]. Because of small sample sizes and proportions near 1, the Freeman–Tukey double arcsine transformation was applied [10]. A random-effects model with REML accounted for between-study variability, and confidence intervals were calculated. P-values for between group comparisons were calculated using multilevel meta regression taking into account the multiple measurements of Steps or LLMs as random intercepts. LLMs struggled with processing the conditional structure of the metadata schema (see Supplemental Figure 1 for details on the schema) which was not conveyed in the prompts. Consequently, data levels were often formatted as data item names and vice versa. For evaluation, output was interpreted as data levels were possible, counting as correct suggestions. Suggestions were also classified as correct if the required data level triggering a certain data item was not selected for a superordinated data item. These implied data entries, however, were not subsequently added to satisfy the schema’s inner logic and, consequently, did not contribute to the count of correct suggestions.

## Results

Across six PILOT articles [11–16], the three‐step pipeline successfully identified all datasets per paper and produced schema‐compliant annotations using repository pages. All articles contained at least one data set with a link to a repository; the maximum number of linked data sets per article was 4. In detail, six datasets were available on the Gene Expression Omnibus (GEO) repository, 3 datasets were available in the European Nucleotide Archive (ENA), one data set was deposited in the Proteomics IDEmAS (PRIDE) database and one in the NCBI Sequence Read Archive (SRA). Last but not least, one set of processed data was published alongside analytical code on GitHub. See Supplemental Figure 2 for details on all datasets used in the interviews.

The outcomes of step 2 are presented in Figure 1. In this step, we restricted extraction to repository records only. Precision, defined as the share of correct annotations among all suggestions, ranged from 82–97%, indicating that the vast majority of predicted biomedical entities were considered correct in the face-to-face interviews. Gemini 2.5 Pro was associated with the highest precision 97.1% (95% CI 94.9%–98.9%), while Claude Sonnet 4 (p<0.001) and GPT-4.1 (p<0.001) performed worse. The mean number of correct entity annotations (expressed as 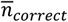 in Figure 1) was 20.2 for Gemini 2.5 Pro,25.5 for GPT-4.1 and 17.7 for Claude Sonnet 4.

**Figure 1:**
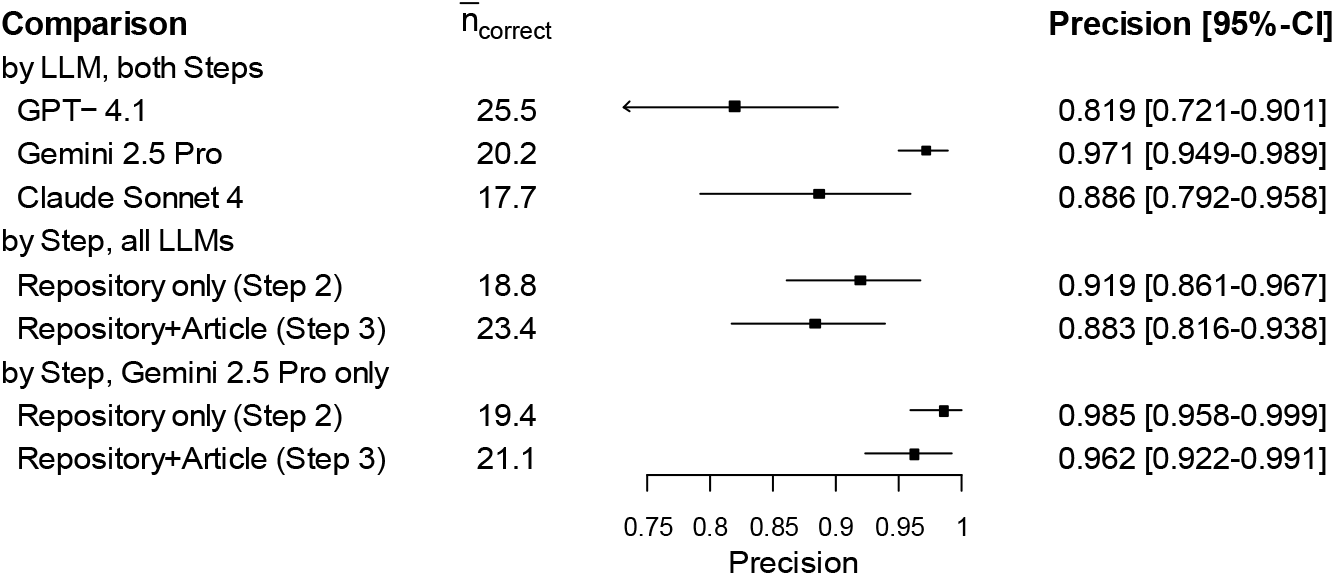
Identification of biomedical entities from multiple repositories using the PILOT metadata schema and search-augmented large language models Step 2 refers to limiting the annotation to the information available in the repositories. Step 3 refers to the situation in which LLMs can use information from the article for metadata enrichment in addition to the repositories. 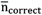 denotes the mean number of annotations classified as correct using face-to-face interviews between a research data management expert and the article’s senior author.

In addition, we found that limiting the annotation to the information available in the repositories achieved better results than adding information from the article. Across all LLMs, the precision was particularly higher in step 2 than in step 3 (91.9% vs 88.3%, p=0.004). At the same time, however the number of correct entity annotations increases when adding information from the article (see Figure 1). When focusing of the results of the best-performing LLM (Gemini 2.5 Pro), the differences in the precision become less profound (98.5% vs. 96.2%, p=0.105). Overall, the results suggest that the restriction to information from online repositories provide a good basis for LLM-supported metadata annotation.

## Discussion

This study demonstrates that search‐augmented, schema‐guided LLM pipelines can reliably discover and annotate multiple datasets per article when grounded in repository landing pages. Relative to article‐augmented extraction, repository‐only extraction improved precision—consistent with the intuition that repository records are curated, canonical source documents, whereas manuscript prose may include shorthand, future‐tense plans, or contextual statements that are easy for LLMs to overgeneralize. Viewed through a FAIR and AI-ready lens, repository-grounded extraction serves as a provenance spine to which model cards, prompts, training/evaluation artifacts, and compute context can be attached for robust reuse [17].

Our results extend earlier grounded‐LLM work on biomedical entity identification by (1) explicitly handling multi‐repository, multi‐dataset publications; (2) formalizing provenance‐aware field filling; and(3) providing a reusable prompt pack aligned to a biomedical metadata schema [6, 7].

The evaluation has several limitations. First, the sample size and scope were restricted to articles within the PILOT context, so generalizability to other domains and repositories remains to be tested at scale. Second, while expert review was used to create reference labels, residual subjectivity is still possible. Third, large language model outputs are inherently variable—non-deterministic and version-dependent—so exact replication requires pinning model versions and prompts. Fourth, the study prioritized precision over recall. This is due to the practical challenges to identify biomedical entities that were not predicted by the LLMs. In natural language processing research, false negatives are often considered in metrics such as accuracy, recall, and the F1 score; however, in this context, the open-ended nature of potential annotations makes such an approach infeasible. While, in principle, missing entities could be identified through author interviews, this would require substantial additional effort. Nevertheless, by guiding the LLM with a predefined CRC metadata schema, the approach ensures that generated annotations are not only contextually appropriate but also relevant to the CRC framework. Lastbut not least, repository heterogeneity presents challenges, as field names and levels of detail vary across repositories, and some schema fields may be under-specified on landing pages.

Implementation of automated dataset annotation would have to respect the conditional inner logic of the metadata schema. Future research will have to address integration of this logic into the machine prompts and devise procedures to ensure compliant data population.

## Data Availability

All data produced in the present study are available upon reasonable request to the authors

---

**Supplemental Figure 1:**
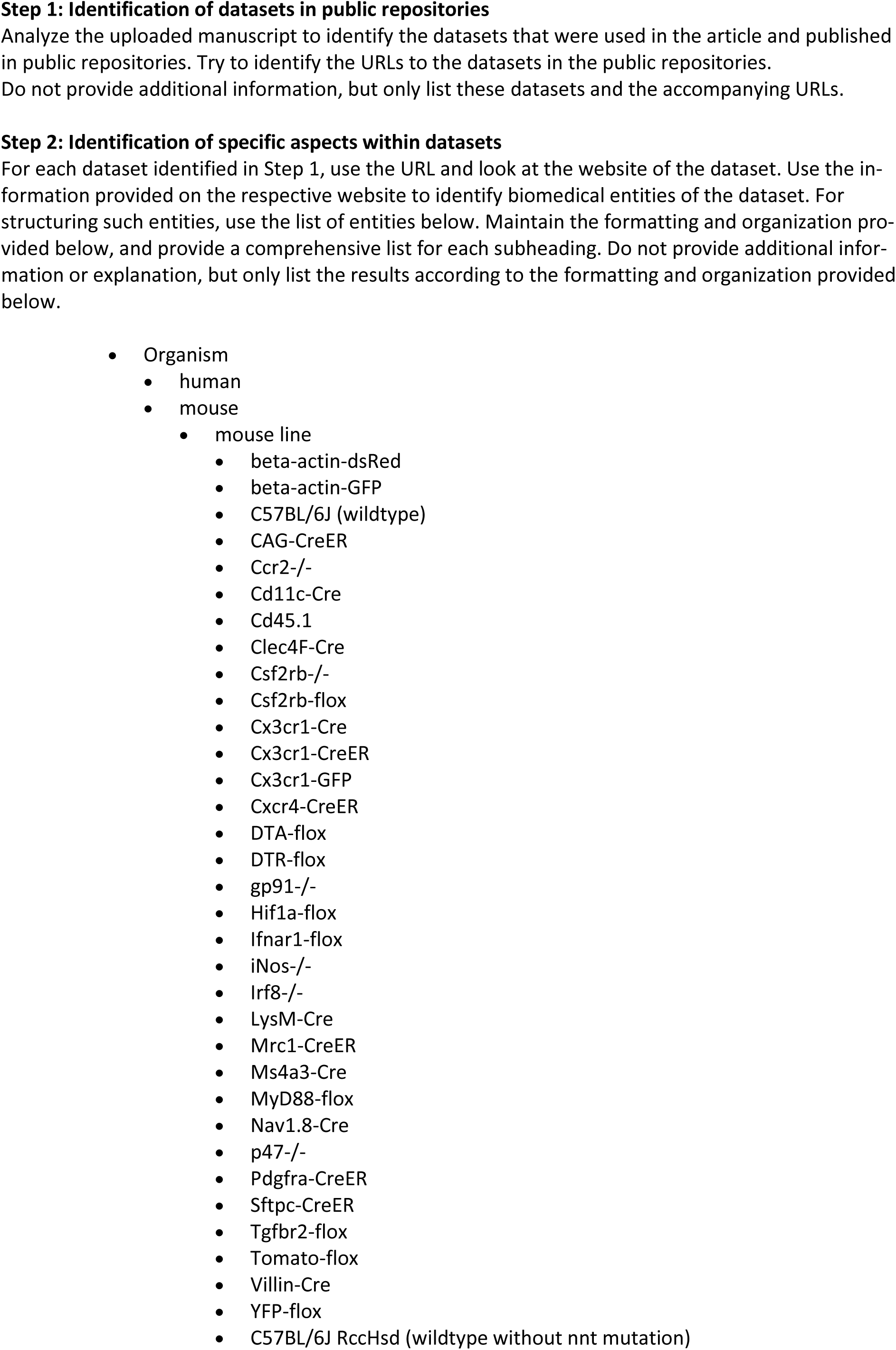
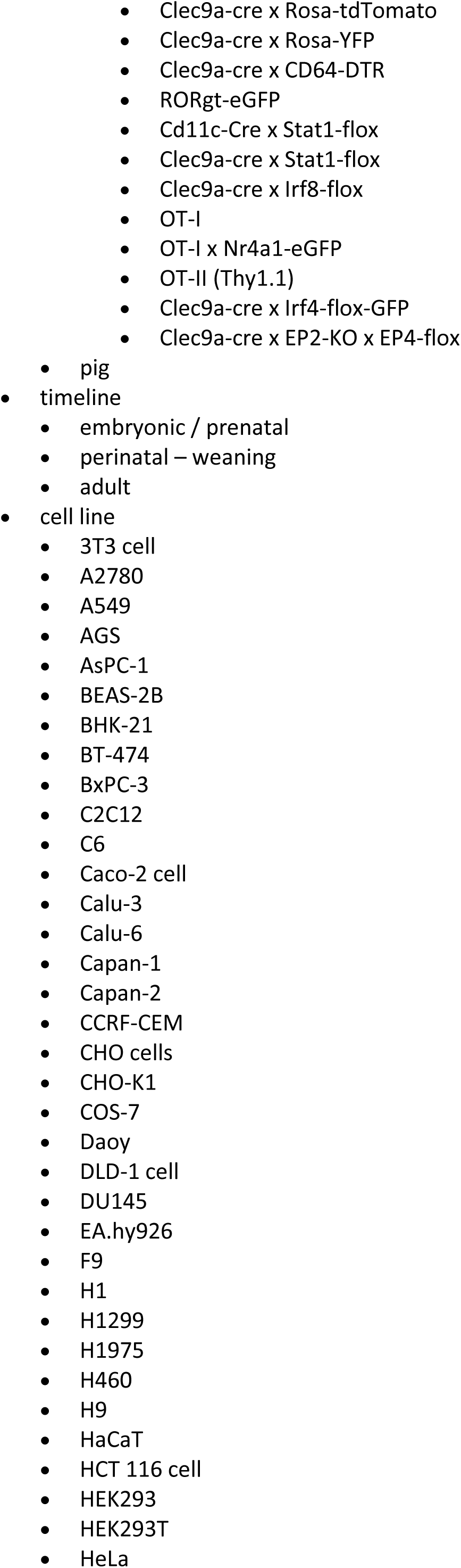
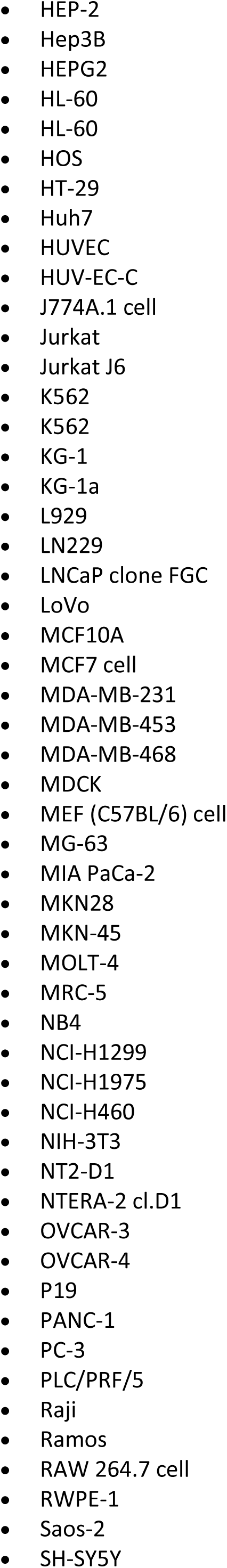
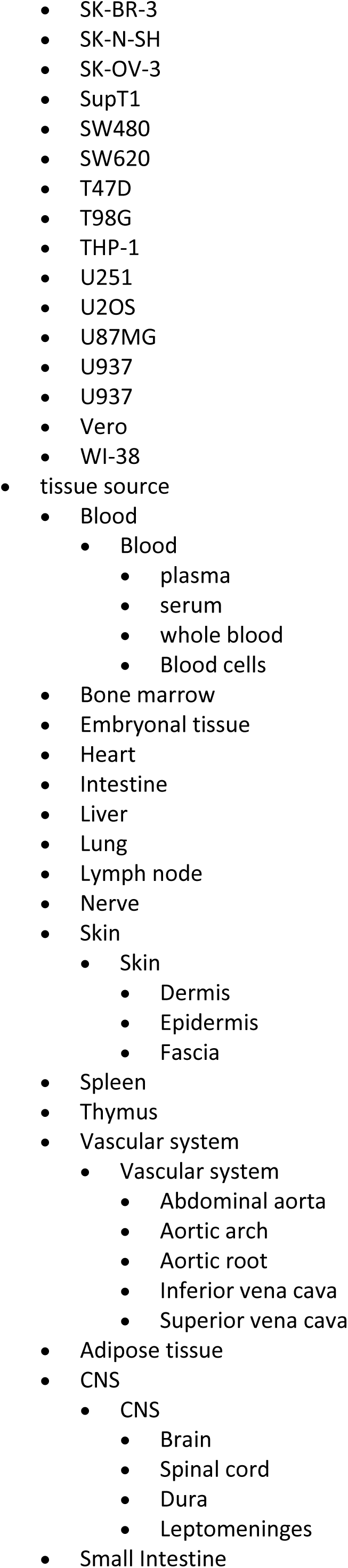
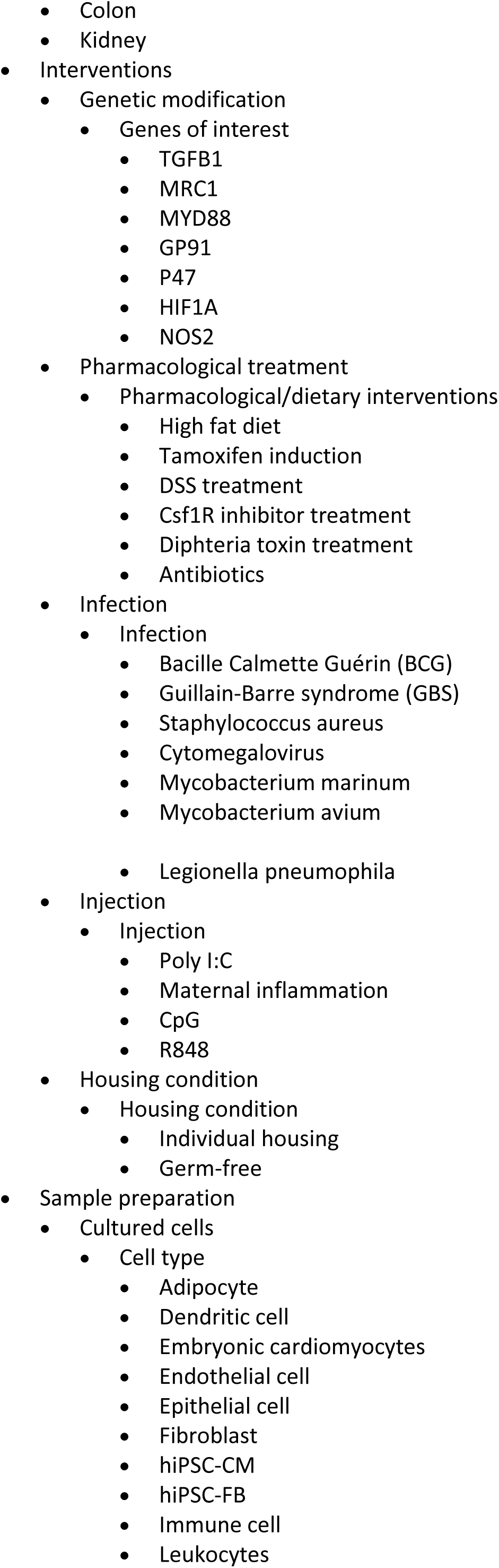
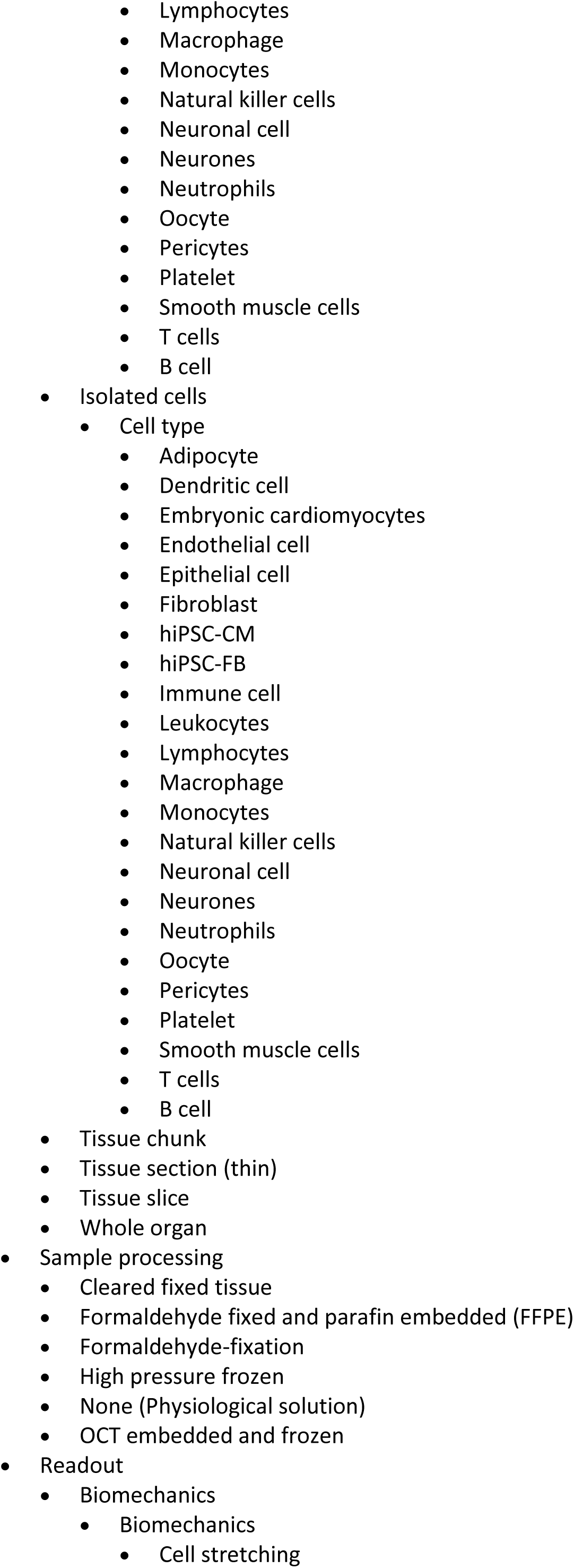
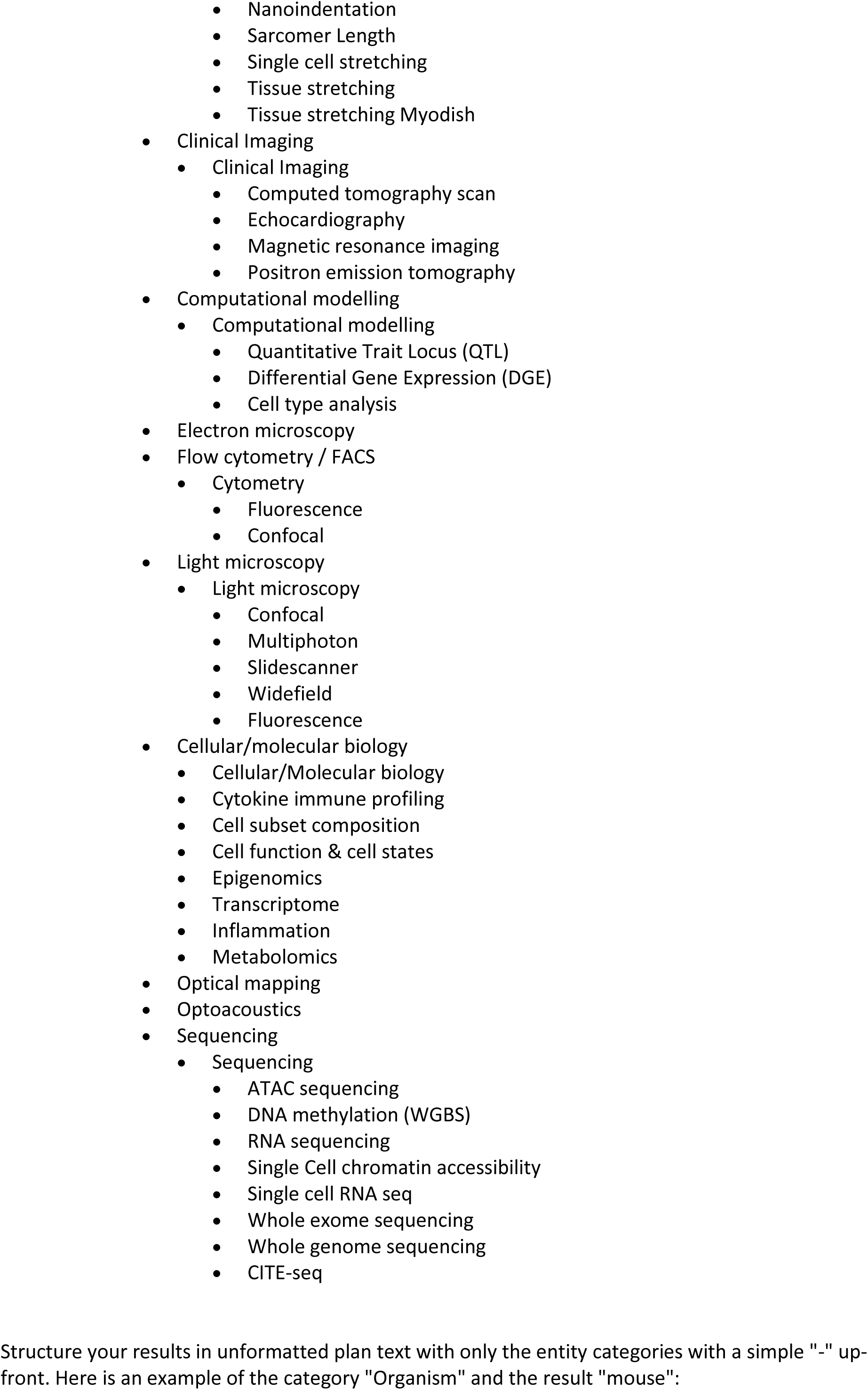
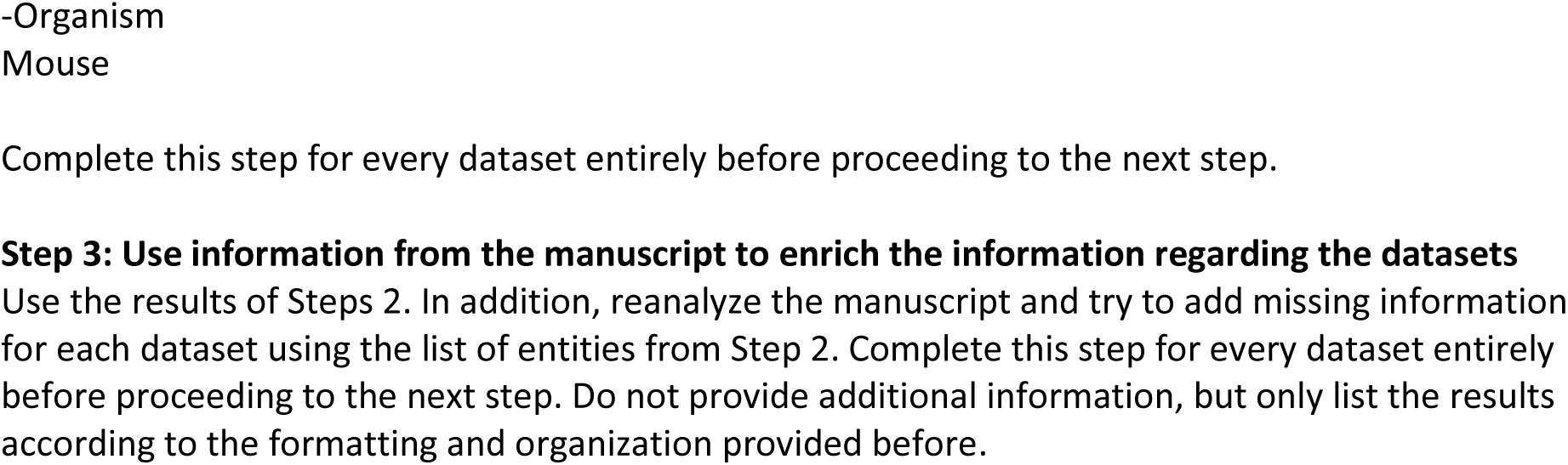
Details on the prompt and schema.

**Supplemental Figure 2:**
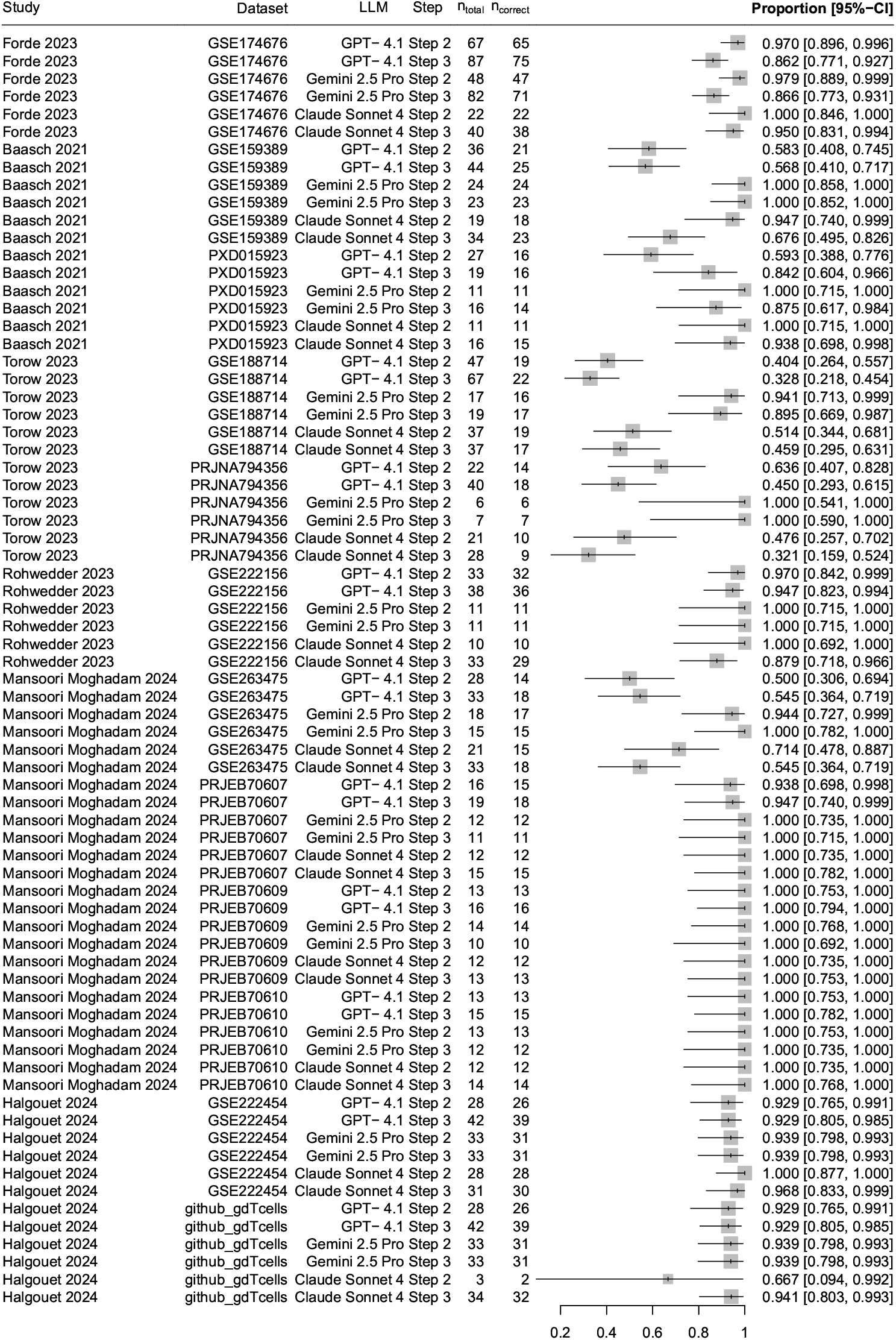
Results of all meta analyses. **Note:** github_gdTcells refers to a datset published at https://github.com/sagar161286/multimodal_gdTcells/

